# TLFT: Transfer Learning and Fourier Transform for ECG Classification

**DOI:** 10.1101/2024.07.09.24310152

**Authors:** Erick Wang, Sarah Lee

## Abstract

Electrocardiogram (ECG) provides a non-invasive method for identifying cardiac issues, particularly arrhythmias or irregular heartbeats. In recent years, the fields of artificial intelligence and machine learning have made significant inroads into various healthcare applications, including the development of arrhythmia classifiers using deep learning techniques. However, a persistent challenge in this domain is the limited availability of large, well-annotated ECG datasets, which are crucial for building and evaluating robust machine learning models. To address this limitation, we propose a novel deep transfer learning framework designed to perform effectively on small training datasets. Our approach involves fine-tuning ResNet-18, a general-purpose image classifier, using the MIT-BIH arrhythmia dataset. This method aims to leverage the power of transfer learning to overcome the constraints of limited data availability. Furthermore, this paper conducts a critical examination of existing deep learning models in the field of ECG analysis. Our investigation reveals that many of these models suffer from methodological flaws, particularly in terms of data leakage. This issue potentially leads to overly optimistic performance estimates and raises concerns about the reliability and generalizability of these models in real-world clinical applications. By addressing these challenges, our work contributes to the advancement of more robust and reliable ECG analysis techniques, potentially improving the accuracy and applicability of automated arrhythmia detection in clinical settings.

## INTRODUCTION

Electrocardiogram (ECG) serves as a crucial non-invasive tool for detecting cardiac abnormalities, particularly arrhythmias or irregular heartbeats. While arrhythmias can occur in healthy individuals, they may also indicate serious cardiac conditions. The traditional method of manually analyzing ECG signals for arrhythmia detection is not only time-consuming but also prone to errors ^1,2^. Recent advancements in deep learning have revolutionized automatic ECG-based arrhythmia diagnosis ^1–14^. However, these methods typically require substantial amounts of training data. Given the scarcity of well-annotated ECG data for arrhythmia detection ^9^, transfer learning techniques utilizing pre-trained image classifiers have gained traction. ^5^ showed that transfer learning is effective when dataset is small. Recent studies have explored transfer learning approaches with the MIT-BIH dataset for developing arrhythmia diagnosis models ^1^. The MIT-BIH arrhythmia database remains the most widely used resource for developing and evaluating ECG-based arrhythmia models ^3,7^.

ECG analysis typically involves four main steps: ECG signal preprocessing and noise attenuation, heartbeat segmentation, feature extraction, and learning/classification ^2^. While the first three steps have been extensively studied in the literature ^10–15^, this section provides a brief overview of selected methods due to space constraints. For ECG signal preprocessing, researchers have proposed various techniques. Sayadi et al. developed a modified extended Kalman filter structure, which serves the dual purpose of denoising and compressing ECG signals ^10^. In the realm of heartbeat segmentation, Li et al. presented a wavelet transform-based algorithm that has shown promise in detecting QRS complexes, even in the presence of high P or T waves and significant noise or drift ^11^. Feature extraction methods have also seen significant development. Lin et al. proposed an automatic heartbeat classification system that utilizes normalized RR intervals and morphological features derived from wavelet transform and linear prediction modeling ^15^. Machine learning models have been widely adopted for arrhythmia classification ^2,3,6,8,9,14–16^. These range from support vector machines with reduced features derived by linear discriminant analysis ^6^ to Hidden Markov Models (HMMs) that combine temporal information and statistical knowledge of ECG signals ^16^. More recently, Hannun et al. developed an end-to-end deep learning approach which directly processes raw ECG signals to produce classifications without the need for feature engineering or selection ^9^. Mousavi and Afghah explored sequence-to-sequence deep learning methods to automatically extract temporal and statistical features from ECG signals ^17^.

We propose a novel end-to-end ECG classification framework that leverages transfer learning. Our approach employs the Fourier Transform (FT) to convert 1D ECG signals into 2D time-frequency domain data, enabling the use of pre-trained 2D CNN models such as VGGs and ResNets. The key contributions of our work are:

1. Development of an end-to-end ECG classification framework that harnesses the power of existing pre-trained 2D CNN models.
2. Exposition of unreliable and biased model evaluation practices in current ECG classification literature using deep learning methods.

## MATERIALS AND METHODS

### Dataset

This study develops the model on the MIT-BIH Arrhythmia dataset ^7^. The dataset includes 48 half-hour excerpts of two-channel ambulatory ECG recordings collected from 47 patients at 360 Hz. The dataset was annotated at heartbeat level by two or more cardiologists independently. 14 original heartbeat types are consolidated into 5 groups. Table ^1^ shows the heartbeat distribution by classes of the raw data, intra-patient split, and inter-patient split. The dataset can be divided into training and testing sets using two distinct approaches: the inter-patient paradigm and the intra-patient paradigm. The intra-patient approach involves randomly selecting heartbeat samples to create the training and testing datasets. This method, however, presents a significant limitation: heartbeat samples from the same patient may appear in both the training and testing sets. Consequently, the testing data could inadvertently influence the model’s training process, potentially leading to overfitting and inflated performance metrics. We posit that this intra-patient data split paradigm can yield unreliable and potentially misleading results. The performance of models developed using this approach may not generalize well to new, unseen patients in real-world clinical scenarios. As such, we strongly advise against the use of the intra-patient approach in ECG classification studies ^4,18^. Furthermore, we recommend that models previously developed and evaluated using the intra-patient split paradigm should be critically reassessed. Their reported performance metrics should be interpreted with caution, and these models should undergo rigorous re-evaluation using more appropriate data splitting techniques before being considered for clinical decision-making applications. In light of these concerns, we advocate for the adoption of the inter-patient paradigm as the standard approach for developing and evaluating ECG classification models. This method ensures a more robust and clinically relevant assessment of model performance, better reflecting the real-world scenario where models must generalize to new, unseen patients.

### Methodology

In our proposed approach, we use pretrained 2D CNN models (ResNet18) which requires the input data to be in the format of 2D images. Therefore, Fourier Transform (FT) is used to obtain 2D time-frequency spectrograms of the digitized 1D ECG recordings for capturing the frequency variations ^14,19^. The 2D time-frequency spectrograms for each point in the signal is computed by ^14^:

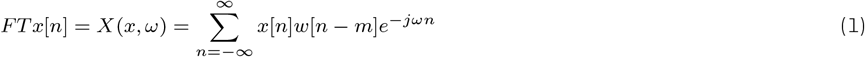

Where *x*[*n*] is the signal which is sampled at 360 Hz and *w*[*n−m*] is the moving window (e.g., Hanning window or Gaussian window). We utilized ResNet18 to classify ECG recordings into four classes. The input data dimensions were adjusted for compatibility with ResNet18. A fully connected layer at the end of ResNet18 was modified to predict the four classes. To classify Arrhythmia, the pretrained ResNet18 network was fine-tuned using the preprocessed training dataset from the inter-patient split paradigm. The retrained ResNet18 was then evaluated using the test dataset. The training parameters for the transfer learning-based model are as follows: Adam optimizer, batch size of 500, training for up to 20 epochs, and a learning rate of 0.0001. The evaluation metrics are precision, recall, and accuracy.

## RESULTS

Table 2 shows that the proposed ResNet18 model with data augmentation achieves the best overall accuracy, the best recall in the normal (N) class, and the best precisions in the arrhythmia (S, V, F) classes.

**Table 1:** Heartbeat distribution by classes of the raw data, intra-patient split, and inter-patient split.

| Set | N | S | V | F | Q | Total |
| --- | --- | --- | --- | --- | --- | --- |
| Full MIT-BIH set | 90,631 | 2,781 | 7,236 | 803 | 8,043 | 109,494 |
| Intra-patient split |  |  |  |  |  |  |
| Training (80% split) | 72,471 | 2,223 | 5,789 | 642 | 6,431 | 87,756 |
| Testing (20% split) | 18,118 | 556 | 1,447 | 161 | 1,608 | 21,890 |
| Inter-patient split |  |  |  |  |  |  |
| Training | 45,866 | 944 | 3,788 | 415 | 8 | 51,021 |
| Testing | 44,259 | 1,837 | 3,221 | 388 | 7 | 49,712 |

**Table 2:** Performance comparison of deep learning models with inter-patient split paradigm. The metrics reported are overall accuracy, precision (Pre), and recall (Rec). Note that the first model was not tested using inter-patient split paradigm in the original paper. The results obtained here are from our re-implementations. The best scores are bold-faced in each column.

| Accuracy (%) | N (n=44,259)<br>Pre/Rec | S (n=1,837)<br>Pre/Rec | V (n=3,221)<br>Pre/Rec | F (n=388)<br>Pre/Rec | Q (n=7)<br>Pre/Rec |
| --- | --- | --- | --- | --- | --- |
| **87.8** | **90.3/91.1** | **45.1/59.0** | **71.2/74.4** | **65.3/70.9** | **83.3/90.9** |

## DISCUSSION & CONCLUSION

We propose an end-to-end ECG classification framework using 2D CNN classifiers. By transforming the 1D ECG waveforms into 2D frequency-time spectrograms using Fourier Transform, this framework allows for the integration of general-purpose pre-trained 2D CNN models (e.g., VGG-16, EfficientNet, etc.) for arrhythmia detection. To build robust and unbiased arrhythmia classifiers, we highly recommend practitioners to follow the correct practice of splitting the training and testing data to avoid any possible information leakage. Moreover, we call for more transparency in data preprocessing and model development, along with the establishment of a standard for model evaluation. This approach will enable the research community to reproduce and verify results effectively.

## Data Availability

All data produced in the present work are contained in the manuscript

